# Burden of intestinal parasitic infections in children and its association with hand washing practice in Ethiopia: a systematic review and meta-analysis

**DOI:** 10.1101/2020.12.12.20248105

**Authors:** Fasil Wagnew, Aster Tadesse, Amanuel Abajobir

**Author notes:** corresponding author: Fasil Wagnew, FW. Email addresses AT, AA.

## Abstract

**Background:** Intestinal parasitic infections (IPIs) are a major public health challenges in developing countries including Ethiopia, although few studies previously estimated the magnitude of IPIs and associated factors in the country. Reports from these scarce studies were also widely varied and remained inconsistent. This study thus aimed to synthesize the pooled magnitude of IPIs and factors affecting it.

**Methods:** Internationally broad based medical database including MEDLINE/PubMed, EMBASE, PsychINFO and Web of Science, and Google Scholar for grey literature were exhaustively searched using *a priori* set criteria to identify studies estimating the prevalence of IPIs among children from 2000-2018. PRISMA guideline was used to systematically review and meta-analyze these studies. Details of study characteristics including sample size, magnitude of effect sizes (including odds ratios (ORs)) and standard errors were extracted. Random-effects model was used to calculate pooled estimates in Stata/se version-14. I^2^ and meta-bias statistics assessed heterogeneity and Egger’s test for publication bias. Sub-group analyses were also carried out based on age of children and regions.

**Results:** Forty-three studies were included in the final analysis (N = 20,008 children). The overall prevalence of IPIs, with one or more species, was 48.2% (95% CI: 40.1, 56.3) in Ethiopian children. Based on sub-group analyses, the highest prevalence of IPIs was observed among school-age children (52.4% (95% CI, 41.3, 63.5)) and in Amhara regional state (52.1% (95% CI, 37.3-66.8)). The odds of having IPIs was nearly six times higher in children who were not practicing hand washing as compared to their counterparts (pooled OR = 5.6 (95% CI: 3.4,9.3). Funnel plot analysis and Egger’s test detected no publication bias.

**Conclusion:** On aggregate, the pooled prevalence of IPIs among Ethiopian children is significantly high. Not hand washing before eating was a risk factor for IPIs. The establishment of applicable sanitation services and health education will help reduce the magnitude of IPIs and promote a healthier childhood.

## Background

Intestinal parasitic infections (IPIs) are serious public health challenges globally [1] whereby one-third of the total population is infected with intestinal parasites (IP), and an estimated 4.5 billion people are at risk of infection with one of the three common IPIs – ascaris lumbricoides (800–1000 million*), hookworm* (700-900 million) and *trichuris trichiura (*500 million) [*2-4*] contributing for significant illnesses and disability-adjusted life year lost [5]. Sadly, around 12% of global burden of disease caused by IPIs occur among school-age children in developing countries [7] especially in sub-Saharan African countries ranging from 27.7% to 95% [8-12], and attributable to major causes of morbidity and mortality among these children [1].

Multifactorial causes contribute to the escalation of IPIs in children including overcrowding, lack of safe water, poor personal hygiene, poor hand washing and nutritional disorders [20, 21]. Indeed, IPIs can be consequences of poor socio-economic status and also contribute to a substantial economic burden [6]. That is, IPIs are closely interrelated with poor socioeconomic status of the family, lack of sanitation, inadequate access to pure water and waste disposal especially in low- and middle-income countries [13-15]. In particular, under-five and school-aged children in these settings are categorized as high-risk population for IPIs [16]. Furthermore, these infections are causes and consequences for anemia and other comorbidities as well as physical and cognitive developmental delays [17, 18] resulting in suboptimal child survival, poor growth and development as well as meager educational performance [8, 10, 18]. In addition, chronic forms of IPIs are associated with the transmission and severity of other infections and/or diseases such as leprosy and human immune-deficiency virus (HIV) [19].

Interestingly, these infections have recently been receiving a high priority from global community. Cognizant to this, the World Health Organization (WHO) has permitted preventive chemoprophylaxis as worldwide mainstay strategy to control the communicability of these infections [22]. In this strategy, scheduled administration of safe and efficacious deworming drugs for risk groups is recommended, setting a minimum target at 75%, and maximum 100% for success rate [22-24]. In fact, general cleanliness or sanitation are imperative indicators to achieve these standards at a community level [25].

Numerous studies have studied the magnitude of IPIs in preschool and school-age children in Ethiopia. However, there is a significant variability in the reports and an aggregate figure is implicated to know the ‘real’ burden of IPIs. We thus carried out a systematic review and meta-analysis on the prevalence of IPIs and its association with common indicators including hand washing practice in children. These analyses were carried out in the context of in-depth methodological application and rigorous statistical analyses that could be helpful for estimating national level burden of IPIs as previous studies are characterized by high spatial variation.

## Methods

### Study design and search strategy

Using broad-based computerized international databases, we employed systematic searches to find all published studies on the prevalence of IPIs in Ethiopian children from 2000-2020. Databases included MEDLINE/PubMed, EMBASE, Psych INFO and Web of Science, and Google Scholar for grey literature. Reference lists of eligible articles, hand searches for grey literature and relevant scientific conference websites were also appraised to enhance electronic searches. Observational studies conducted on the prevalence of IPIs among children in Ethiopia were selected for this systematic review and meta-analysis. Search protocol was developed by using such keywords as “prevalence” OR “Epidemiology” AND “intestinal parasite,” “intestinal helminthes”, “intestinal protozoa”, “soil-transmitted helminthes” AND “associated factors” OR “predictors” AND “Ethiopia”. To make a certain scientific rigor, we strictly adhered to the Preferred Reporting of Systematic Reviews and Meta-Analysis (PRISMA) guideline [26].

### Inclusion criteria

All studies written in English, estimating the prevalence of IPIs in Ethiopian children and of high-quality scores were entered into the final meta-analysis.

### Exclusion Criteria

Studies that did not estimate the prevalence of IPIs, articles without definite sample size or full-text, those that did not achieve the minimum quality scores, case reports, case series, and experimental studies were excluded from the meta-analysis.

### Data Extraction

Two reviewers (FW and AT) screened the downloaded titles and abstracts using the eligibility criteria. Discussions and mutual consensus were in place when possible arguments raised between the reviewers. Two reviewers (FW and AT) then assessed full-texts of potentially eligible papers. We made some efforts to communicate the authors whenever further information was needed. Numerator and denominator data and beta coefficients and their standard errors (if given) were used to calculate ORs, where ORs with 95% CI were not available. The data extraction format included first author, study year, region of study, study design and sample size, and prevalence of IPIs.

### Quality Appraisal

Articles were assessed for quality, with only high-quality studies included in the analysis. Two authors (FW, AT) independently evaluated the quality of each relevant article. The reviewer’s cross-checked their quality appraisal scores and resolved any disparity before calculating the final appraisal score. Moreover, Kappa statistic was used to measure agreement between reviewers to determine uniformity of amongst eligible full-text articles using guidelines proposed by Landis and Koch [31]: < 0.20 = slight agreement; 0.21–0.40 = fair agreement; 0.41–0.60 = moderate agreement; 0.61–0.80 = substantial agreement; and > 0.80 = almost perfect agreement. Finally, AA reviewed overall quality of extracted data and edited the manuscript. Newcastle-Ottawa Scale adapted for prevalence studies methodological quality assessment tool was also used [27]. The tool has three components in general–the first component graded as five stars gives a due emphasis on the methodological quality of each original study. The second component of the tool explores the comparability of the study. The third component focuses on the outcome and statistical analysis of studies. Articles with a score of ≥ 6 out of 10 were considered as high-quality. Accordingly, all eligible studies had high quality scores.

### Data analysis

Information about study designs, study sample and country were summarized using Microsoft Excel and then exported to STATA/se version-14 for analysis. Meta-analysis of the magnitude of IPIs was carried out using a random-effects model, generating a pooled prevalence with 95% CIs. Heterogeneity among included studies was assessed using Cochran’s Q and I^2^ statistic [28]. I^2^ statistic estimates the percentage of heterogeneity across studies. Forest plots were also used to estimate the pool effect sizes and weight of overall and each included study with respective CI to offer a graphic summary of the data. Publication bias was determined based on the symmetry of funnel plots [29], and Egger’s test [30]. Sub-group analysis was carried out, based on age of participants and regional states, because of the presence of significant heterogeneity across the included studies.

## Results

Our search yielded 2,896 articles; of these, 442 duplications and 2,411 unrelated articles were excluded. Kappa coefficient revealed that agreement rate between the two reviewers for the included articles was 0.81. Fifteen full-text articles [32-46] were excluded due to unmet outcome of interest or location. Finally, 43 eligible studies representing 20,008 participants met the inclusion criteria for this review that determine the burden of IPIs (**Figure 1)**.

**Figure 1:**
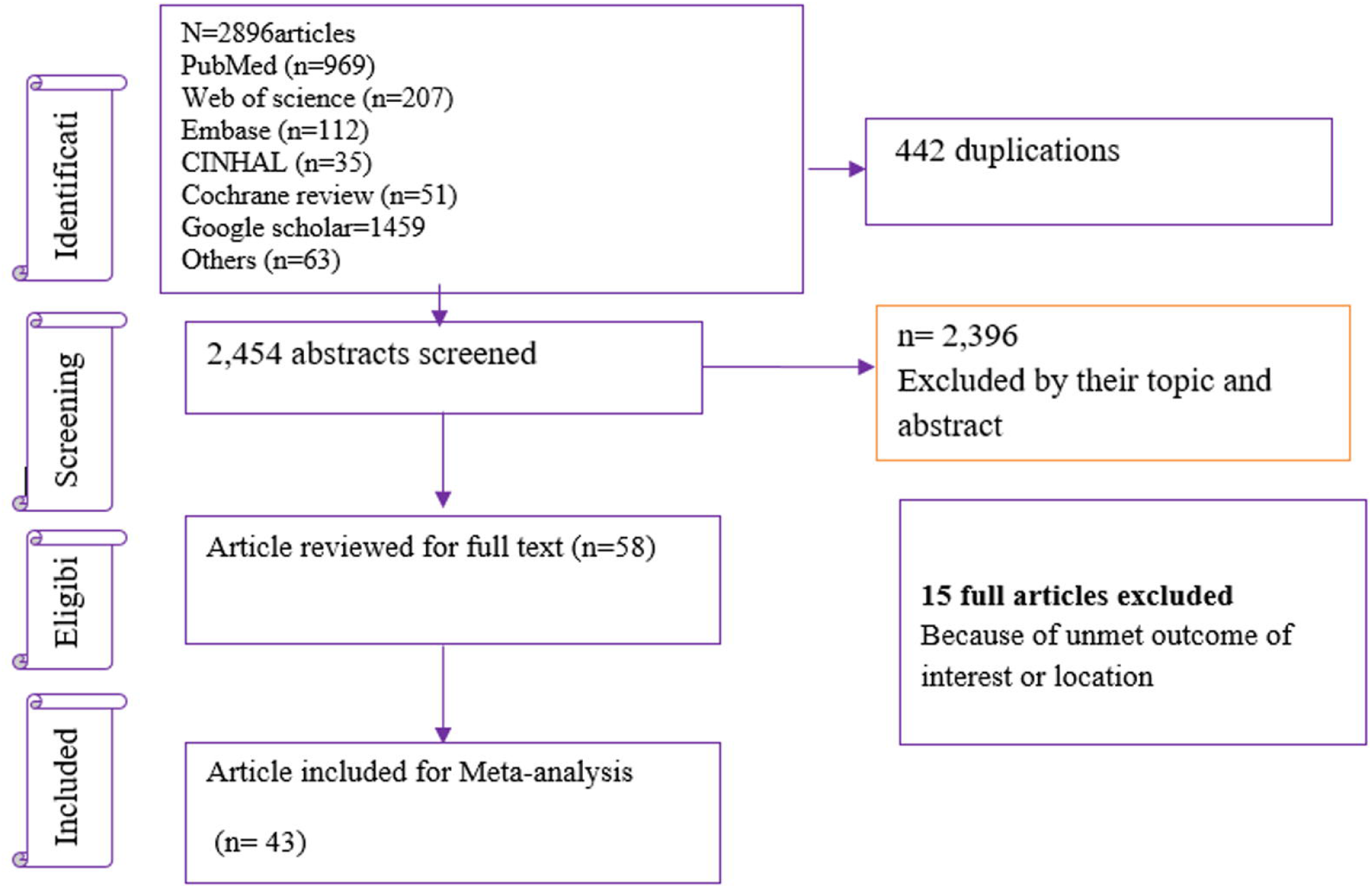
Flowchart diagram describing selection of studies for the burden of IPIs and association with residence and hand washing practice in Ethiopian children, 2018

### Descriptive characteristics of the included studies

Descriptive characteristics for the included epidemiological studies is presented in Table 1. Of these, 42 studies were cross-sectional in design and only one study was case-control [47]. Children were classified based on presence versus absence of an IPI for any species. The largest sample size was reported by the study conducted by Grimes J et al [50] and smallest for the study done by Mulatu G [16]. Similarly, the prevalence of IPI was highest for the study conducted by Worku L et al [48] and lowest for the study conducted by Zemene T [49] (**Table 1**).

**Table 1:**
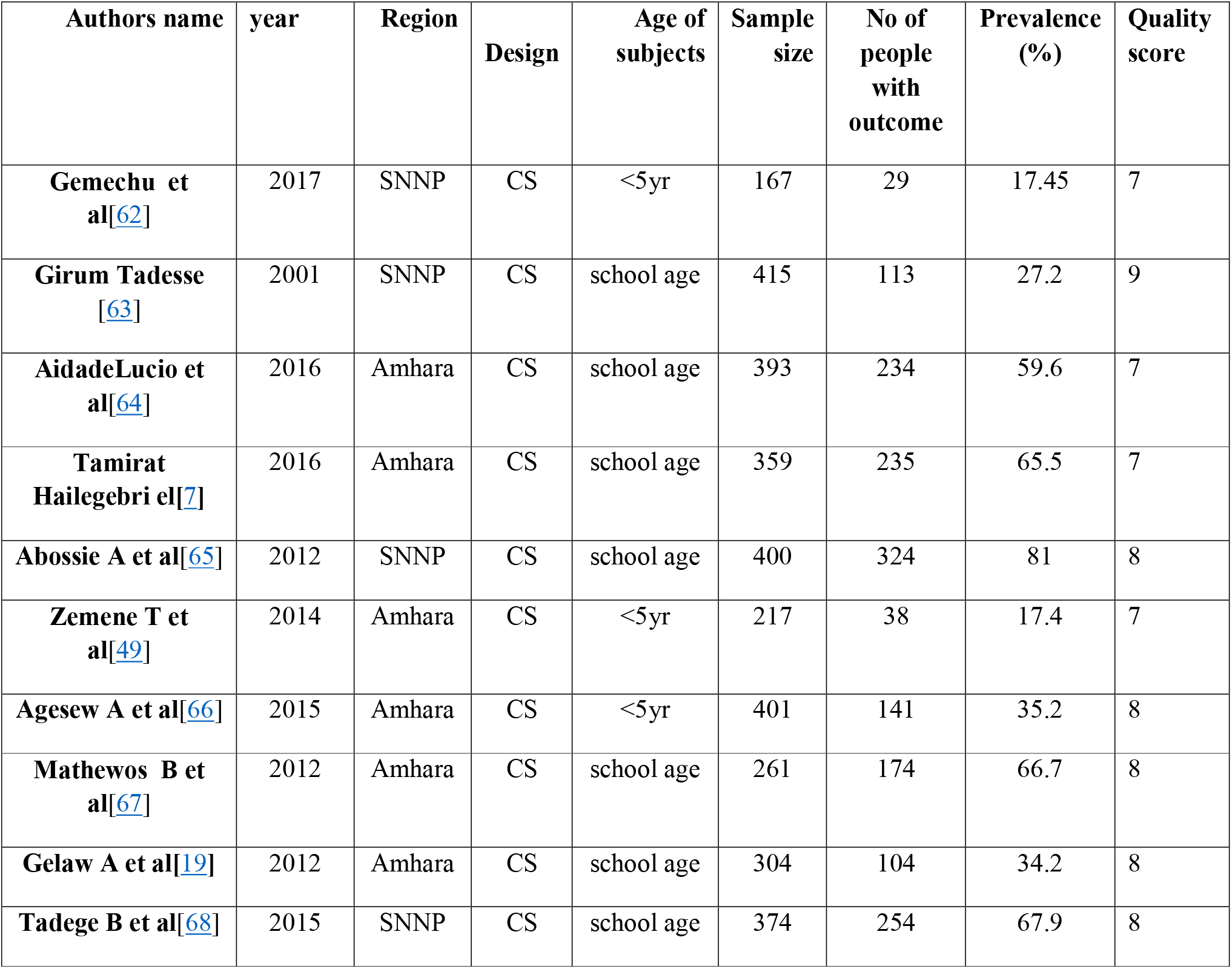
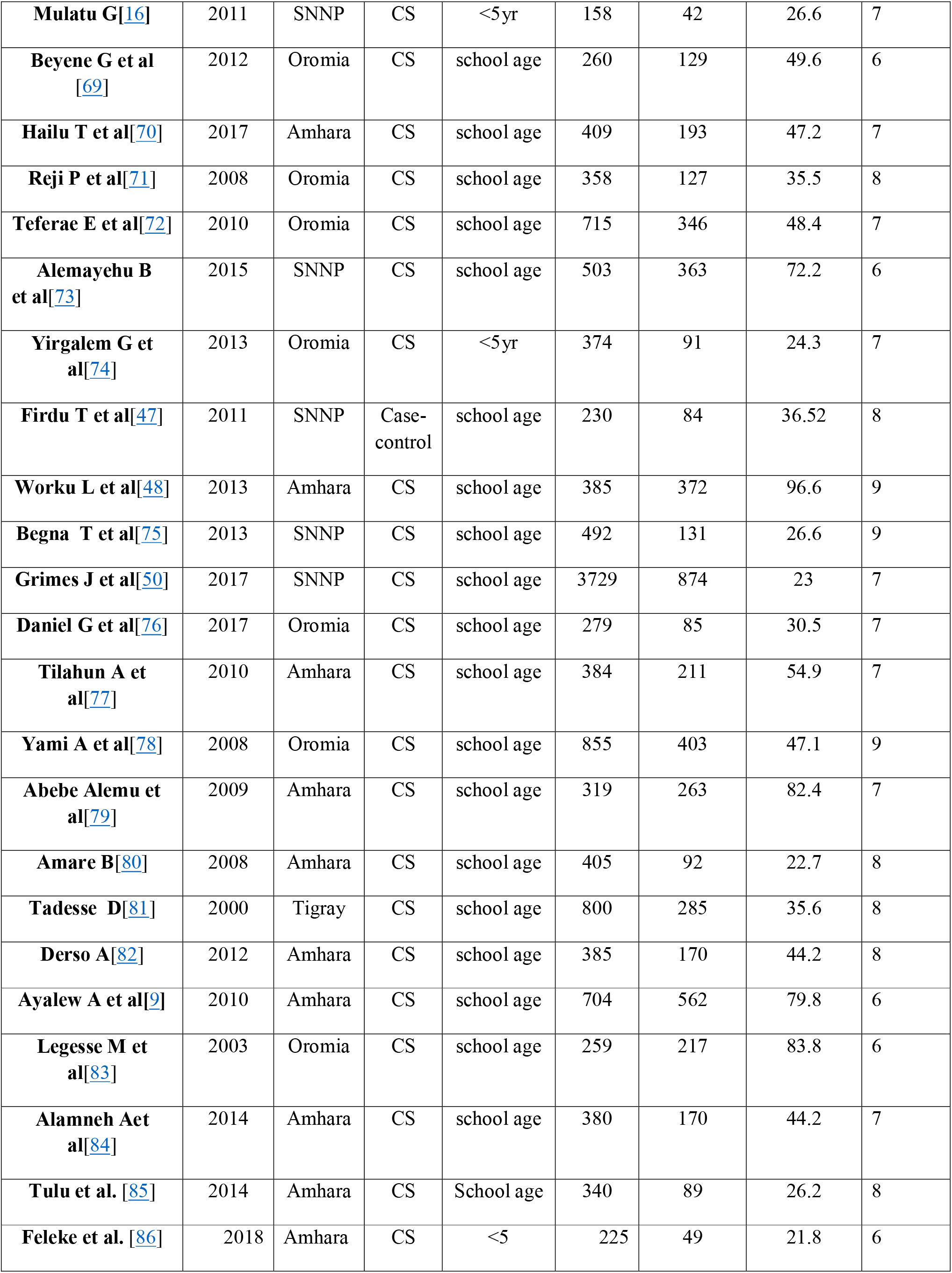
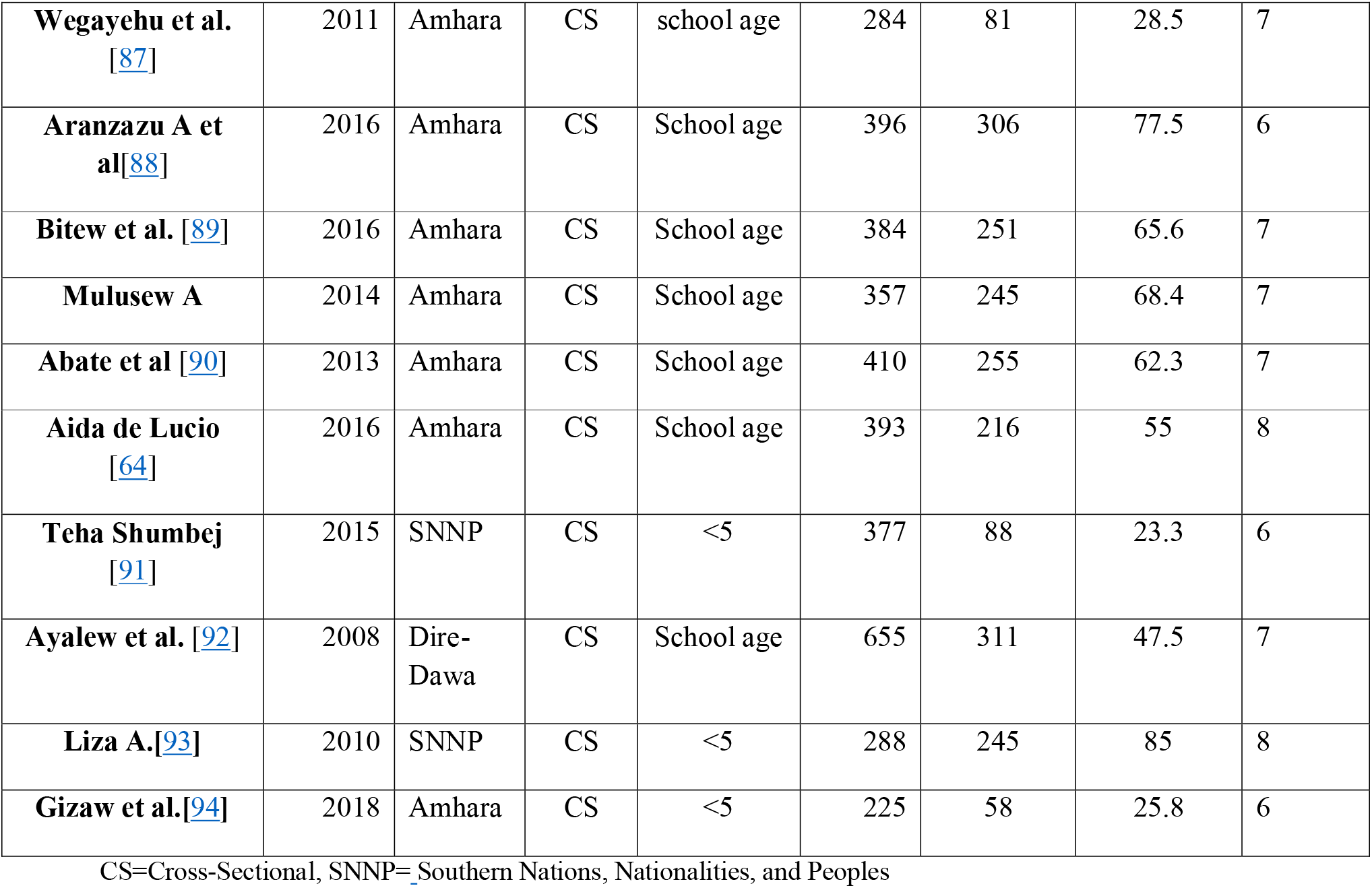
Descriptive summary of 31 included studies on the prevalence of intestinal parasitic infections and association with residence and hand washing in Ethiopian children.

| Authors name | year | Region | Design | Age of subjects | Sample size | No of people with outcome | Prevalence (%) | Quality score |
| --- | --- | --- | --- | --- | --- | --- | --- | --- |
| Gemechu et al[62] | 2017 | SNNP | CS | <5yr | 167 | 29 | 17.45 | 7 |
| Girum Tadesse [63] | 2001 | SNNP | CS | school age | 415 | 113 | 27.2 | 9 |
| AidadeLucio et al[64] | 2016 | Amhara | CS | school age | 393 | 234 | 59.6 | 7 |
| Tamirat Hailegebriel[7] | 2016 | Amhara | CS | school age | 359 | 235 | 65.5 | 7 |
| Abossie A et al[65] | 2012 | SNNP | CS | school age | 400 | 324 | 81 | 8 |
| Zemene T et al[49] | 2014 | Amhara | CS | <5yr | 217 | 38 | 17.4 | 7 |
| Agezew A et al[66] | 2015 | Amhara | CS | <5yr | 401 | 141 | 35.2 | 8 |
| Mathewos B et al[67] | 2012 | Amhara | CS | school age | 261 | 174 | 66.7 | 8 |
| Gelaw A et al[19] | 2012 | Amhara | CS | school age | 304 | 104 | 34.2 | 8 |
| Tadege B et al[68] | 2015 | SNNP | CS | school age | 374 | 254 | 67.9 | 8 |
| **Mulatu G**[16] | 2011 | SNNP | CS | <5yr | 158 | 42 | 26.6 | 7 |
| **Beyene G et al**<br>[69] | 2012 | Oromia | CS | school age | 260 | 129 | 49.6 | 6 |
| **Hailu T et al**[70] | 2017 | Amhara | CS | school age | 409 | 193 | 47.2 | 7 |
| **Reji P et al**[71] | 2008 | Oromia | CS | school age | 358 | 127 | 35.5 | 8 |
| **Teferae E et al**[72] | 2010 | Oromia | CS | school age | 715 | 346 | 48.4 | 7 |
| **Alemayehu B et al**[73] | 2015 | SNNP | CS | school age | 503 | 363 | 72.2 | 6 |
| **Yirgalem G et al**[74] | 2013 | Oromia | CS | <5yr | 374 | 91 | 24.3 | 7 |
| **Firdu T et al**[47] | 2011 | SNNP | Case-control | school age | 230 | 84 | 36.52 | 8 |
| **Worku L et al**[48] | 2013 | Amhara | CS | school age | 385 | 372 | 96.6 | 9 |
| **Begna T et al**[75] | 2013 | SNNP | CS | school age | 492 | 131 | 26.6 | 9 |
| **Grimes J et al**[50] | 2017 | SNNP | CS | school age | 3729 | 874 | 23 | 7 |
| **Daniel G et al**[76] | 2017 | Oromia | CS | school age | 279 | 85 | 30.5 | 7 |
| **Tilahun A et al**[77] | 2010 | Amhara | CS | school age | 384 | 211 | 54.9 | 7 |
| **Yami A et al**[78] | 2008 | Oromia | CS | school age | 855 | 403 | 47.1 | 9 |
| **Abebe Alemu et al**[79] | 2009 | Amhara | CS | school age | 319 | 263 | 82.4 | 7 |
| **Amare B**[80] | 2008 | Amhara | CS | school age | 405 | 92 | 22.7 | 8 |
| **Tadesse D**[81] | 2000 | Tigray | CS | school age | 800 | 285 | 35.6 | 8 |
| **Derso A**[82] | 2012 | Amhara | CS | school age | 385 | 170 | 44.2 | 8 |
| **Ayalew A et al**[9] | 2010 | Amhara | CS | school age | 704 | 562 | 79.8 | 6 |
| **Legesse M et al**[83] | 2003 | Oromia | CS | school age | 259 | 217 | 83.8 | 6 |
| **Alamneh A et al**[84] | 2014 | Amhara | CS | school age | 380 | 170 | 44.2 | 7 |
| **Tulu et al.** [85] | 2014 | Amhara | CS | School age | 340 | 89 | 26.2 | 8 |
| **Feleke et al.** [86] | 2018 | Amhara | CS | <5 | 225 | 49 | 21.8 | 6 |
| **Wegayehu et al. [87]** | 2011 | Amhara | CS | school age | 284 | 81 | 28.5 | 7 |
| **Aranzazu A et al[88]** | 2016 | Amhara | CS | School age | 396 | 306 | 77.5 | 6 |
| **Bitew et al. [89]** | 2016 | Amhara | CS | School age | 384 | 251 | 65.6 | 7 |
| **Mulusew A** | 2014 | Amhara | CS | School age | 357 | 245 | 68.4 | 7 |
| **Abate et al [90]** | 2013 | Amhara | CS | School age | 410 | 255 | 62.3 | 7 |
| **Aida de Lucio [64]** | 2016 | Amhara | CS | School age | 393 | 216 | 55 | 8 |
| **Teha Shumbej [91]** | 2015 | SNNP | CS | <5 | 377 | 88 | 23.3 | 6 |
| **Ayalew et al. [92]** | 2008 | Dire-Dawa | CS | School age | 655 | 311 | 47.5 | 7 |
| **Liza A.[93]** | 2010 | SNNP | CS | <5 | 288 | 245 | 85 | 8 |
| **Gizaw et al.[94]** | 2018 | Amhara | CS | <5 | 225 | 58 | 25.8 | 6 |

CS=Cross-Sectional, SNNP= Southern Nations, Nationalities, and Peoples

### Prevalence of IPIs in children

The overall prevalence of IPIs with one or more species was 48.2% (95% CI: 40.1, 56.3%) in Ethiopian children (**Figure 2**). We performed subgroup analyses based on regional states and age of participants as there was significant hetrogeniety across included studies. For instance, the prevalence of IPIs was 52.4% (95% CI: 41.3, 63.5) in school-age children and 52.1% (95% CI: 37.3, 66.8) of children with IPIs lived in Amhara regional state (**Table 2**). Overall Egger’s test revealed no statistically significant publication bias, p-value = 0.62 (**Additional file 1**).

**Table 2:** Subgroup analysis based on regions and age of participants among diabetes patients

| **Variables** | **Characteristics** | **Estimates with 95% CI** | **P-value** |
| --- | --- | --- | --- |
| **Age of participant** | Under-five children | 24.23(17.44-31.02) | <0.001 |
|  | School-age children | 52.43(41.37-63.5) |  |
| **Region** | Amhara | 52.1(37.3-66.8) | <0.001 |
|  | Oromia | 45.1(30.9-59.3) |  |
|  | SNNP | 43.9(24.9-62.9) |  |
|  | Tigray | 35.6(32.2-38.9) |  |

SNNP= Southern Nations, Nationalities, and Peoples

**Figure 2:**
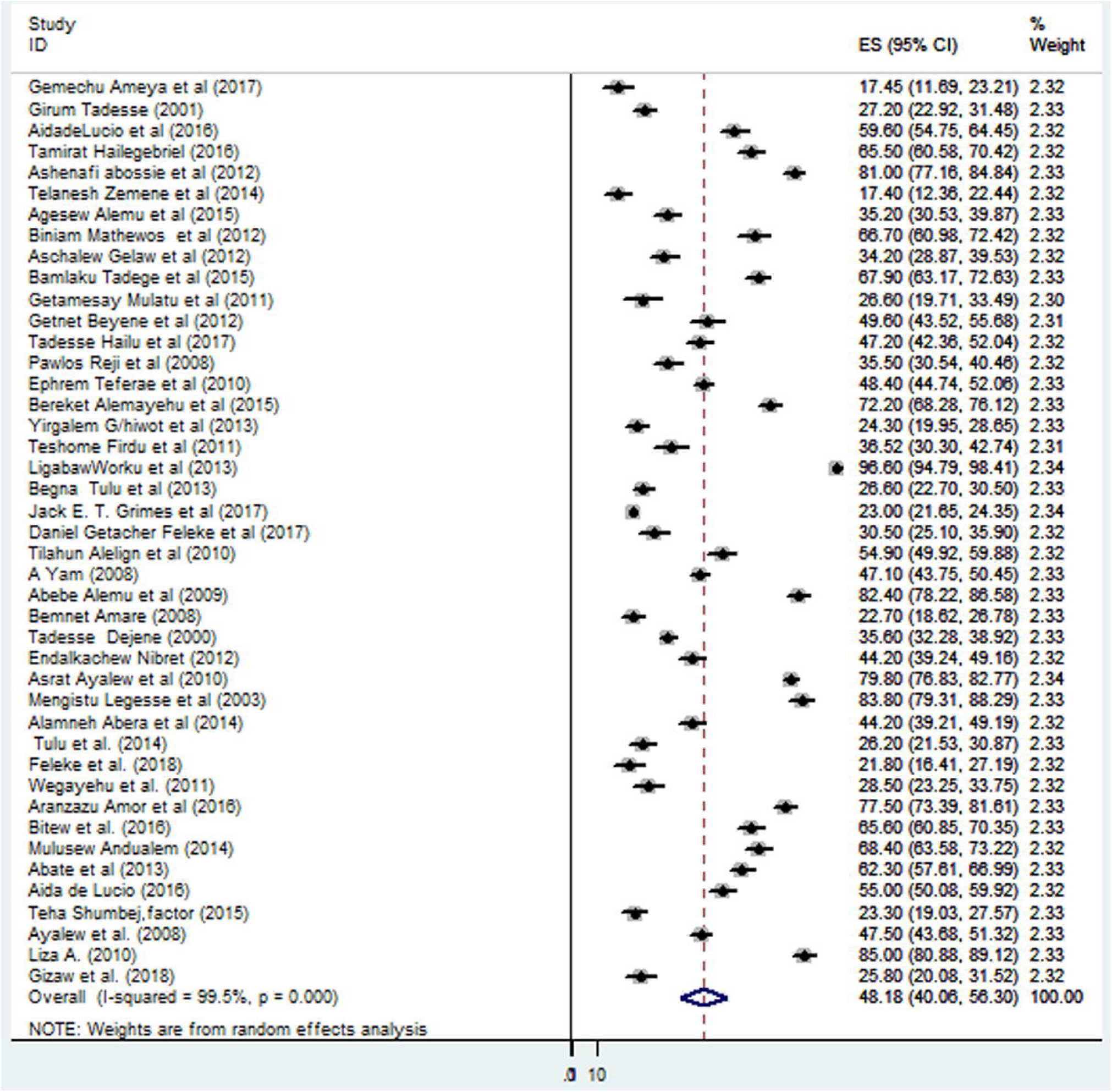
Forest Plot of the 31 studies that quantitatively assessed the pooled prevalence of IPIs, 2018.

### Association of residence and hand washing practice with IPIs

Five studies were included to determine the association of hand washing before eating on IPIs (**Table 3**). Children without hand washing before eating experienced six times more likely the occurrence of IPIs (OR: 5.6 (95% CI: 3.4, 9.3)) as compared to hand washing before eating (**Figure 3**). However, the association between residence (urban versus rural) showed a non-significant association with IPIs (OR: 1.1 (95% CI: 0.5, 2.1) (**Figure 4**).

**Table 3.** the effect of hand washing before eating on intestinal parasitic infections in Ethiopian children.

| Authors and year | Region | Hand washing | <u>Intestinal parasites</u> |  | OR(with 95% CI) |
| --- | --- | --- | --- | --- | --- |
|  |  | Before eating | Positive | negative |  |
| Gemechu A et al,2017[62] | Southern | Yes | 7 | 83 | 1 |
|  |  | NO | 22 | 25 | 4.70(1.88-11.75) |
| Ayalew A et al,2010[9] | Amhara | Yes | 261 | 110 | 1 |
|  |  | No | 301 | 32 | 3.96(2.58-6) |
| Abera A et al,2014[84] | Amhara | Yes | 135 | 191 | 1 |
|  |  | No | 35 | 24 | 2.06(1.17-3.62) |
| Tadege B et al,2015[68] | SNNP | Yes | 57 | 156 | 1 |
|  |  | No | 140 | 21 | 14.02(8-24.3) |
| Alemu A et al,2015[66] | Amhara | Yes | 6 | 96 | 1 |
|  |  | No | 86 | 213 | 6.46(2.73-15.3) |
| Hailu et al, 2017[70] | Amhara | Yes | 128 | 227 | 1 |
|  |  | No | 40 | 14 | 2.90(1.52-5.53) |
| Teha et al. 2015[91] | SNNP | Yes | 67 | 186 | 1 |
|  |  | No | 21 | 191 | 4.4(2.6-7.6) |
| Gizew et al. 2018[94] | Amhara | Yes | 10 | 92 | 1 |
|  |  | No | 48 | 75 | 5.8(2.7-12.4) |
| Tegegne E. et al. 2017[95] | Amhara | Yes | 22 | 148 | 1 |
|  |  | No | 43 | 10 | 28.9(12.7-65.7) |

`SNNP= Southern Nations, Nationalities, and Peoples

**Figure 3:**
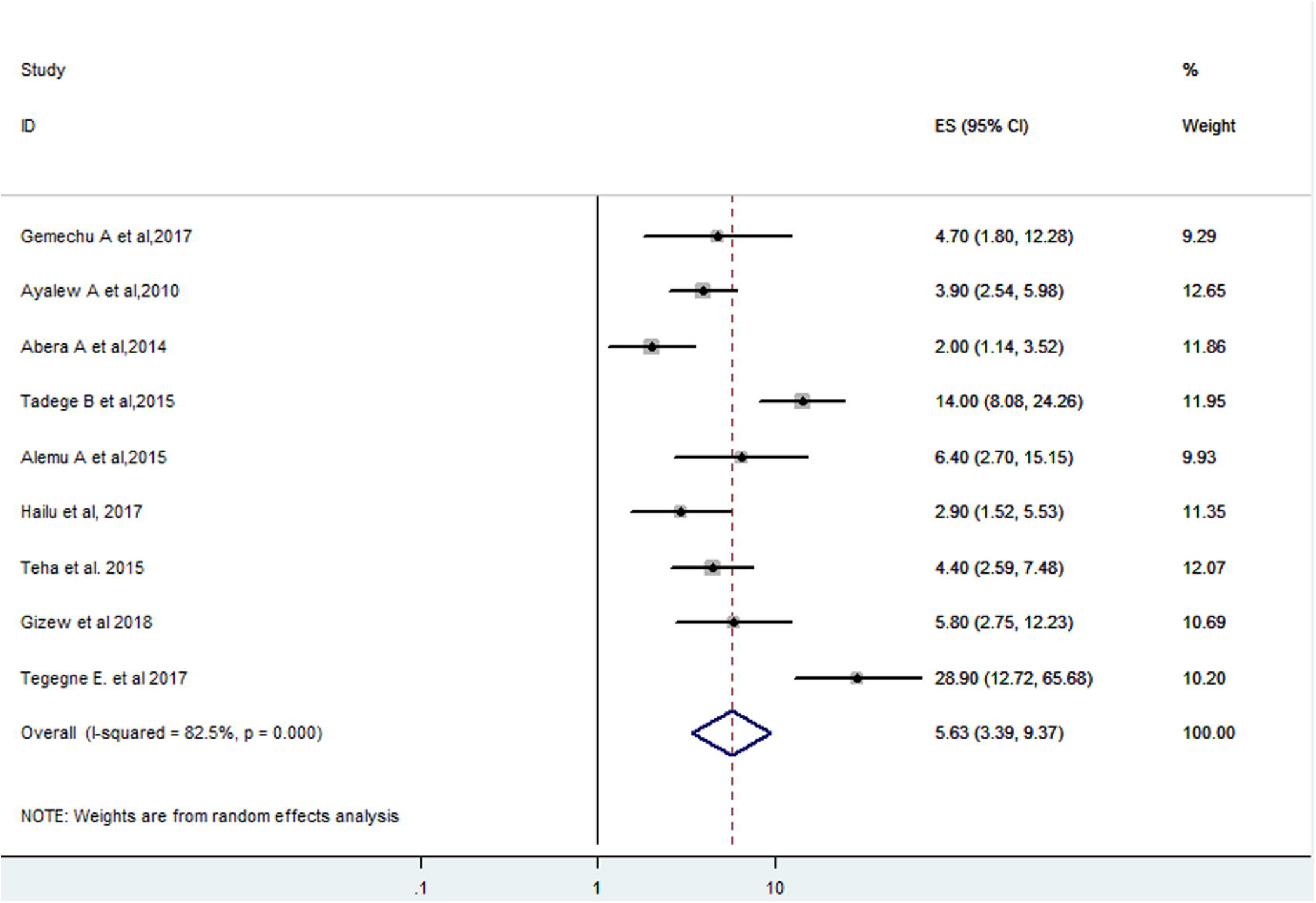
Forest Plot of the 6 studies that quantitatively assessed the effect of hand washing before eating on the occurrences of IPIs, 2018

**Figure 4:**
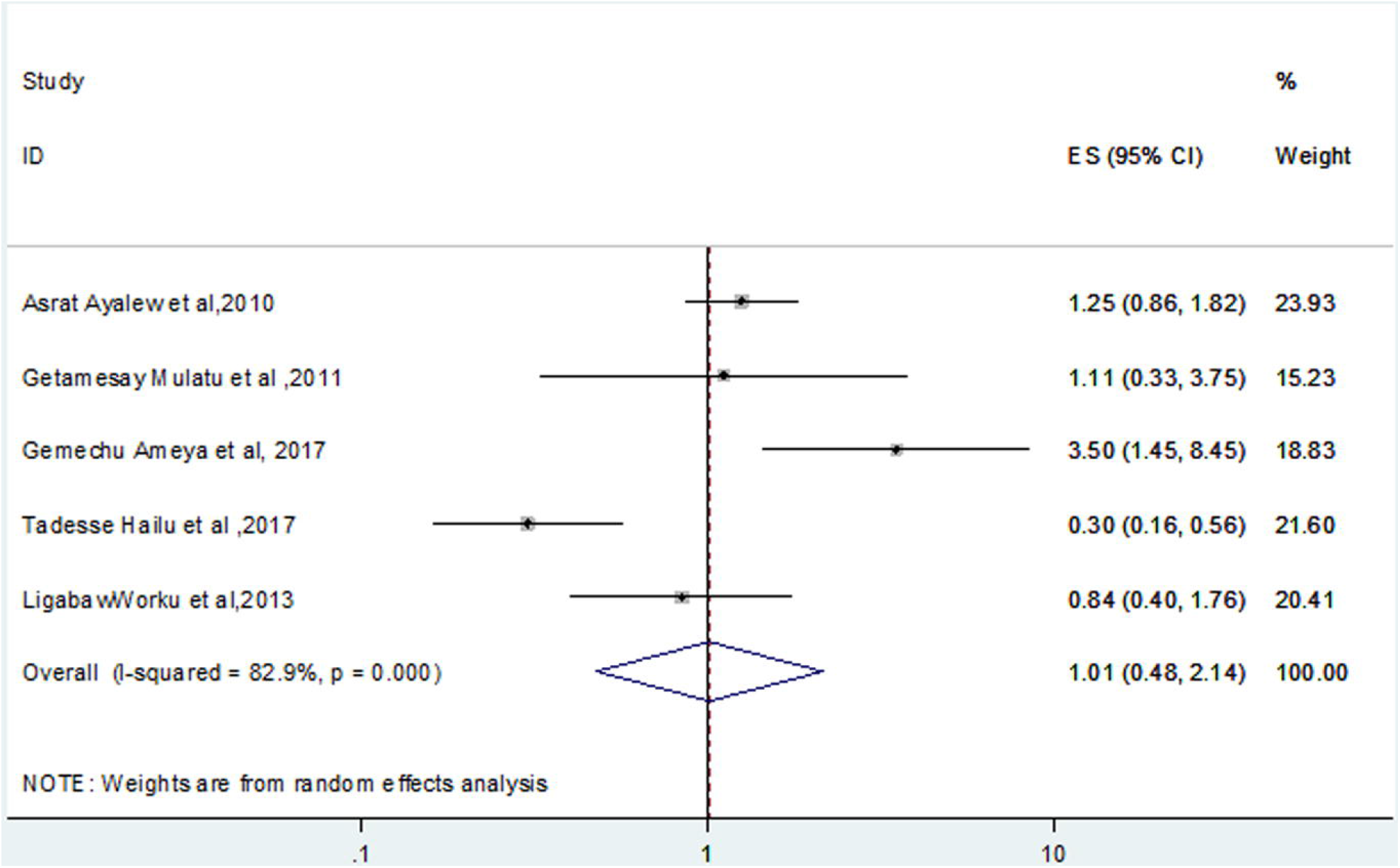
Forest Plot of the 6 studies that quantitatively assessed the effect of residence on the occurrences of IPIs, 2018.

## Discussions

This study estimated the pooled prevalence of IPIs with one or more species in Ethiopian children and its association with residence and hand washing practice, using the documented data from systematic literature review, which was assembled from different regions of the country. After diligent search through scientific databases, scientific reports and grey literature, 43 studies were included in the final analysis. Accordingly, the overall pooled prevalence of IPIs among Ethiopian children was 48.2%. This finding was in line with previous studies that revealed 42.5% IPIs in Syria [51], 47.6% in Afghanistan [52] and 38% in Iran [53]. In contrast, the prevalence of IPIs observed in current study is higher compared to studies in Thailand that reported 38.8% [54], in Shush country (5.14%) [55], in Turkey (17%) [56] and in Korea (10.5%) [57]. This observed variation could be due to socio-economic status, poor hygienic practice and sanitary facilities, inadequate safe drinking water and environmental factors as well as different methods used for the diagnosis of IPIs.

We performed sub-group analysis based on the age of participants and regional states of the country. Accordingly, the evidence revealed that the prevalence of IPIs among school-age children was two-fold higher than that in under five-children. The difference observed between under-five and school-age children might be because of the following reasons–first, close contact between students and unhygienic conditions in the school compound; second, variation in sanitation and human development indices; and finally, due to implementation of continuing deworming programme for under-five children by ministry of health. Likewise, the highest prevalence of IPIs were observed in Amhara regional states and the lowest prevalence was in Tigray state. This regional discrepancy might be because of cultural habits (sanitary status) among the regions, variations in methodology and sample size. The advancement of public health measures and policy implementation could also vary across regions that might attribute for the difference.

In an effort to examine the association between residence and hand washing before eating and IPIs, hand washing before eating was associated with a significant reduction in the occurrence of IPIs. Indeed, IPIs is frequently caused by *hookworms, ascaris lumbricoides* and *trichuris trichiura* and affected over one billion individuals globally [58]. This could be due to probable to be hand contaminated with cysts of *giardia, cryptosporidium* and other species. The meager association between residence and IPIs in this meta-analysis is supported by previous studies [60, 61]. This might be because of variations among studies, although Egger’s test for publication bias showed no statistically significant evidence, p-value = 0.06.

The findings of this study may have both clinical and public health implications towards scaling up effective and efficient management of IPIs by informing decision makers and practitioners. In other words, understanding the magnitude and distribution of IPIs is an important precondition for planning and evaluating intervention programs. For instance, safe water and personal hygiene as a basic human right will indisputably result in major health improvements and prevention of different diseases including parasitic infections [59]. That is, since almost all of IPIs are feco-oral communicable infections, delivery of tape water and latrines, health education on personal hygiene and environmental sanitation are crucial to control and reduce IPIs in the country.

However, these findings need to be considered in the context of some important limitations; these include: (i) significant heterogeneity across studies might have contributed for meager association between residence and IPIs, although presumably children in urban areas have better access to personal and environmental hygiene; (ii) this study did not determine other possible risk factors contributing to the occurrence of IPIs; (iii) the gold standard diagnosis of Pinworm infection is Gram staining, though several studies had employed only wet smear; and (iv) stool samples were taken at once but standard diagnosis usually take at least 3 samples.

## Conclusion

This meta-analysis summarized a high pooled prevalence of IPIs in Ethiopian children especially among school-age children. Establishment of applicable sanitation services and health education for hand washing will help make a healthier childhood.

## Supporting information

Suplemental Flow diagram

## Data Availability

The datasets used and/or analyzed during the current study are available from the corresponding author on reasonable request.

## List of abbreviations

AA: Amanuel Abajobir
AT: Aster Tadesse
FW: Fasil Wagnew
IPIs: Intestinal Parasitic Infections
OR: Odds Ratio

## Declarations

### Ethics approval and consent to participate

Not applicable

### Consent to publication

Not applicable

### Availability of data and material

- The datasets used and/or analyzed during the current study are available from the corresponding author on reasonable request.
- PRISMA checklist

## Authors’ contribution

FW involved in the conception of research idea; (FW, AT, AA) undertook data extraction, analysis, interpretation, and manuscript write-up. All authors read and approved the final manuscript.

## Acknowledgements

The authors would like to acknowledge the Debre Markos University library for providing us with a wide range of available online databases.

## Additional Files

**Additional file 1:-Funnel plot of the included studies for checking publication bias**.

