## Supplementary material for "Burden of intestinal parasitic infections in children and its association with hand washing practice in Ethiopia: a systematic review and meta-analysis": Suplemental Flow diagram

**PRISMA Flow Diagram**

**Screening**

**Included**

**Eligibility**

**Identification**

Records identified through database searching
(n = 2,833)

Additional records identified through other sources
(n = 63)

Records after duplicates removed
(n =2,454)

Records screened
(n =2,454)

Records excluded
(n =2,396)

Full-text articles assessed for eligibility
(n = 58)

Full-text articles excluded, with reasons
(n =15)

Studies included in qualitative synthesis
(n = 43)

Studies included in quantitative synthesis (meta-analysis)
(n = 43)
